# Bayesian network modelling for early diagnosis and prediction of Endometriosis

**DOI:** 10.1101/2020.11.04.20225946

**Authors:** Rachel Collins, Norman Fenton

## Abstract

Bayesian networks (BNs) are graphical models that can combine knowledge with data to represent the causal probabilistic relationships between a set of variables and provide insight into the processes underlying disease progression, closely resembling clinical decision-making. This paper describes a BN causal model for the early diagnosis and prediction of endometriosis. The causal structure of the BN model is developed using an idioms-based approach and the model parameters are derived from the data reported in multiple medical observational studies and trials. The BN incorporates the impact of errors and omissions in reporting endometriosis, and the distinction between assumed and actual cases. Hence, it is also able to explain both flawed and counterintuitive results of observational studies.

## Introduction

Endometriosis is a relatively common and potentially debilitating gynaecological disorder that affects 6-10% of women of reproductive age (Rogers, et al., 2013). It is present in about a quarter of women who are infertile (Chen, et al., 2019). Endometriosis is a long-term condition defined as the presence of endometrial-like tissue outside the uterus and it is associated with a chronic and inflammatory reaction. Symptoms of endometriosis can include dysmenorrhoea (painful periods), dyspareunia (pain experienced during intercourse), non-cyclical pelvic pain and subfertility (Farquhar, 2000). The standard diagnosis is based on visualisation and histological examination of the lesions. The wide variety of symptoms of endometriosis often leads to wandering and medical diagnostic delay; it can take an average of four to eleven years for diagnosis with severity of symptoms and probability of diagnosis increasing with age (Akter, et al., 2019; Bérubé, et al., 1998). The cause for delay of diagnosis and treatment may be due to invasive laparoscopic procedures alongside histologic confirmation being the gold standard diagnostic confirmation of the condition and physicians treating the symptoms rather than the condition. It is estimated that affected individuals lose 10.8 hours of work weekly due to this condition and report a decreased quality of life (Rogers, et al., 2013). A better understanding and approach to diagnose this condition would allow better management of these patients.

This paper intends to assess risk and prediction of endometriosis in a Bayesian causal model approach, by building a Bayesian Network (BN) and structuring it with use of medical idioms elicited from expert knowledge and data available, with the aim of improving upon the current statistical models and aid decision-making for professionals. It will primarily function as a diagnostic model for the prediction of suspected endometriosis and probability of an individual being formally diagnosed. It will also offer insight into the impact endometriosis has on the quality of life of the individual.

## Literature Review

### Endometriosis and Machine Learning

The use of both supervised and unsupervised machine learning has been applied to biology studies and the prediction of endometriosis (Tarca, et al., 2007; Wölfler, et al., 2005). Biological patterns have been analysed by various machine learning tools; decision tree, partial least squares discriminant analysis (PLSDA), support vector machine, and random forest to classify endometriosis (Akter, et al., 2019; Lu, et al., 2015). Machine learning techniques have been utilised to explore associations between persistent organic pollutants and deep endometriosis (Matta, et al., 2020). However, this author found limited studies focusing on the signs and symptoms, most often observed in a primary care setting, associated with endometriosis as predictors for diagnosis. Furthermore, this author found Bayesian Networks had been applied as cancer prediction tools but were not utilised for the prediction of Endometriosis as a condition (Reijnen, et al., 2020; Roy, et al., 2015).

Literature was reviewed which claimed to be using Bayesian approaches but were not centred on the concept of casual Bayesian Network models. Correlated modelling framework to estimate ROC curves and the associated area under the curves was applied to the diagnostic performance of physicians in their diagnosing of endometriosis which suggested that clinical information may play a more important role in physicians’ diagnostic performance than their experiences (Chen, et al., 2019). Endometriosis phenotypes were learned from patient-generated data through unsupervised learning (Urteaga, et al., 2020). Bayesian network meta-analyses were used to assess the role traditional Chinese medicine has in the treatment of endometriosis (Dong, et al., 2019).

The use of big data and machine learning algorithms, in which there is the belief of the ability to discover all properties and relationships for prediction and decision-making, may not be appropriate in the analysis of healthcare due to the limited nature of relevant data, the inconsistency of clinician reporting and recording of symptoms in clinic and hospital settings (Fenton & Neil, 2019). Therefore, the use of BN causal models, which can account for unobserved variables, would be a valuable method to use for the prediction of endometriosis in a clinical setting.

### Bayesian Approach to Endometriosis

It must be noted that the literature reviewed involved case control studies that compare incidence of diagnosis, or risk of endometriosis, and control groups. This data is used to calculate odds ratio and confidence interval. Odds ratio is a measure that describes the odds of a patient developing the condition provided a certain criterion is met. It measures the strength of association between two events:

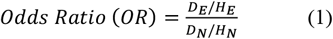

where *D*_*E*_ represents patients who have the condition (endometriosis) and have had a certain sign/symptom, *H*_*E*_ is number of patients who are healthy but have the sign/symptom, *D* _*N*_ the patients who do not have the sign/symptom and do not have the condition, and *H* _*N*_ the patients who are healthy and do not have the sign/symptom. Given an odds ratio, a conditional probability table *B*|*A* can be populated, where *A* can symbolise the sign/symptom and *B* the disease, as shown in Table 1.

**Table 1.** Odds ratio probability table

| | $B = 1$ | $B = 0$ |
| --- | --- | --- |
| $A = 1$ | $p_{11}$ | $p_{10}$ |
| $A = 0$ | $p_{01}$ | $p_{00}$ |

The treatment and use of odds ratios can massively exaggerate the probability of the disease. It is not just potential confounding variables that can compromise the simple ‘odds ratio’ measure of risk factors. These odds ratios do not account for multiple cause and effects or the dependencies of interacting variables. It assumes that the exposure of the variable is the sole explanation for the condition. Yet, in randomised controlled trials (RCTs), often used in medical studies, it is impossible to control for all confounding variables. Odds ratio cannot work for the prediction of endometriosis as the aetiology of the condition is unknown (Farquhar, 2000). By using a Bayesian network for analysis, multiple observations and interventions can be applied and account for potential unobserved data.

### Risk factors associated with Endometriosis

The aetiology of endometriosis has been studied and theorised since 1927 when John A Sampson presented his hypothesis, to the American Gynaecological Society, that the cause was due to retrograde menstruation; the uterine contents reaching the ovaries and the surrounding tissue via a retrograde flow of the menstrual contents through the fallopian tubes (Dastur & Tank, 2010). This theory is now viewed as a sign of endometriosis rather than the cause, as not all women who experience retrograde menstruation develop the condition (Bérubé, et al., 1998). Accordingly, there is general consensus among experts that the cause of endometriosis is still unknown (Bérubé, et al., 1998; Farquhar, 2000; Harris, et al., 2018). As a result, the study of risk factors associated with endometriosis is the focus of many, oftentimes contradictory, studies.

Reports show that there is an increased frequency of the disease among women of higher socio-economic status. Whilst socio-economic status can be viewed an indicator of the lifestyle habits that are associated with an increased risk of endometriosis, such as reproductive pattern, body mass, physical activity and diet (Parazzini, et al., 2017), there is argument that this status, which is closely linked to race and ethnicity, may impact the diagnosis of endometriosis through a patient’s ability to access healthcare (Bougie, et al., 2019).

Diet has often been studied as a potential area of risk for the contraction of endometriosis. There are conflicting studies surrounding the association of consumption of red meat, ham and trans fats with the risk of developing endometriosis (Parazzini, et al., 2013). A 2004 Italian study by Parazzini et al. (2004) reports the risk of endometriosis is significantly higher (OR 2.0, 95% CI 1.4–2.8, Ptrend = 0.0004) for women reporting high meat and ham consumption compared with women reporting a lower intake (OR 1.8, 95% CI 1.3–2.5, Ptrend = 0.001). However, this result was not replicated in a 2011 Trabert et al. (2011) study that summarised that there was no association between endometriosis risk and number of servings per week of red meat. The contradiction indicates further studies are required. Currently, there is no concrete evidence of associations between smoking and caffeine intake and the risk of developing endometriosis (Parazzini, et al., 2017). There is evidence of an association between alcohol consumption and endometriosis risk, but further studies are required to clarify whether alcohol is exacerbating an existing disease or is related to the severity of the disease (Parazzini, et al., 2013).

There is increasing evidence that patients with endometriosis have a higher risk of developing other chronic diseases. Endometriosis patients have a higher risk of ovarian and breast cancers, cutaneous melanoma, asthma, some autoimmune, cardiovascular, and atopic diseases, and a decreased risk of cervical cancer (Kvaskoff, et al., 2014). These findings show why improved screening practices and better care and management for diagnosed patients are needed. Women with endometriosis have an increased risk of Irritable Bowel Syndrome (IBS). There is an association between endometriosis and rheumatoid arthritis and psoriasis which supports the idea that endometriosis includes immunological factors (Parazzini, et al., 2017). Case studies have shown a greater prevalence of endometriosis among women with asthma, allergies, and allergic rhinitis (Kvaskoff, et al., 2014).

Studies do show that predictors of a diagnosis of endometriosis variables reflect the patterns that clinicians observe in a GP setting. A study by Burton et al. (2017) used primary care data to identify patterns of symptoms over time that appeared to be useful pointers to the diagnosis of endometriosis. This study argues that symptoms, such as distinct episodes of gynaecological pain and combinations of gynaecological pain on one occasion with menstrual symptoms or lower gastrointestinal symptoms on another, provide patterns that can be incorporated into diagnostic support systems (Burton, et al., 2017). The presence of endometriosis manifests into specific features of pain, menstrual bleeding, ovarian symptoms including cysts, and subfertility and in the non-specific features of fatigue, gynaecological issues including vulvo-vaginal symptoms and pelvic inflammation, and lower gastro-intestinal issues including pain, bloating and irritable bowel syndrome (Burton, et al., 2017).

During the literature evaluation, it became evident that many studies identifying the ‘risk factors’ associated with endometriosis were, in fact, identifying the signs and symptoms associated with the condition. It became an important focus of this study to separate risk factors from symptoms to correctly apply the direction of influence.

## Methodology

### Bayesian Networks

A causal model is a precise specification of how each variable is influenced by its parents in a Directed Acyclic graph (Pearl, 2009). The model is developed with nodes representing different variables with direct edges representing either a causal, or influential, relationship. For example, in Figure 1 a), Family History is the parent of Suspected Endometriosis, indicating that family history has a causal impact on the probability of Endometriotic disease. In Figure 1 b), the Node Probability Table (NPT) is displayed representing the conditional probability distribution of each node, given the influence of its parent. As Family History is without parents it is referred to as a *root node* and its NPT is the probability distribution of itself (Fenton & Neil, 2019). The Bayesian Network is fully parameterised once all Node Probability tables are supplied. From this, Bayesian probabilistic reasoning, in which the prior belief about a certain hypothesis is updated considering new evidence, can be performed. *Prior probability* is the term used for the initial belief; the updated belief is termed *posterior probability*.

**Figure 1.**
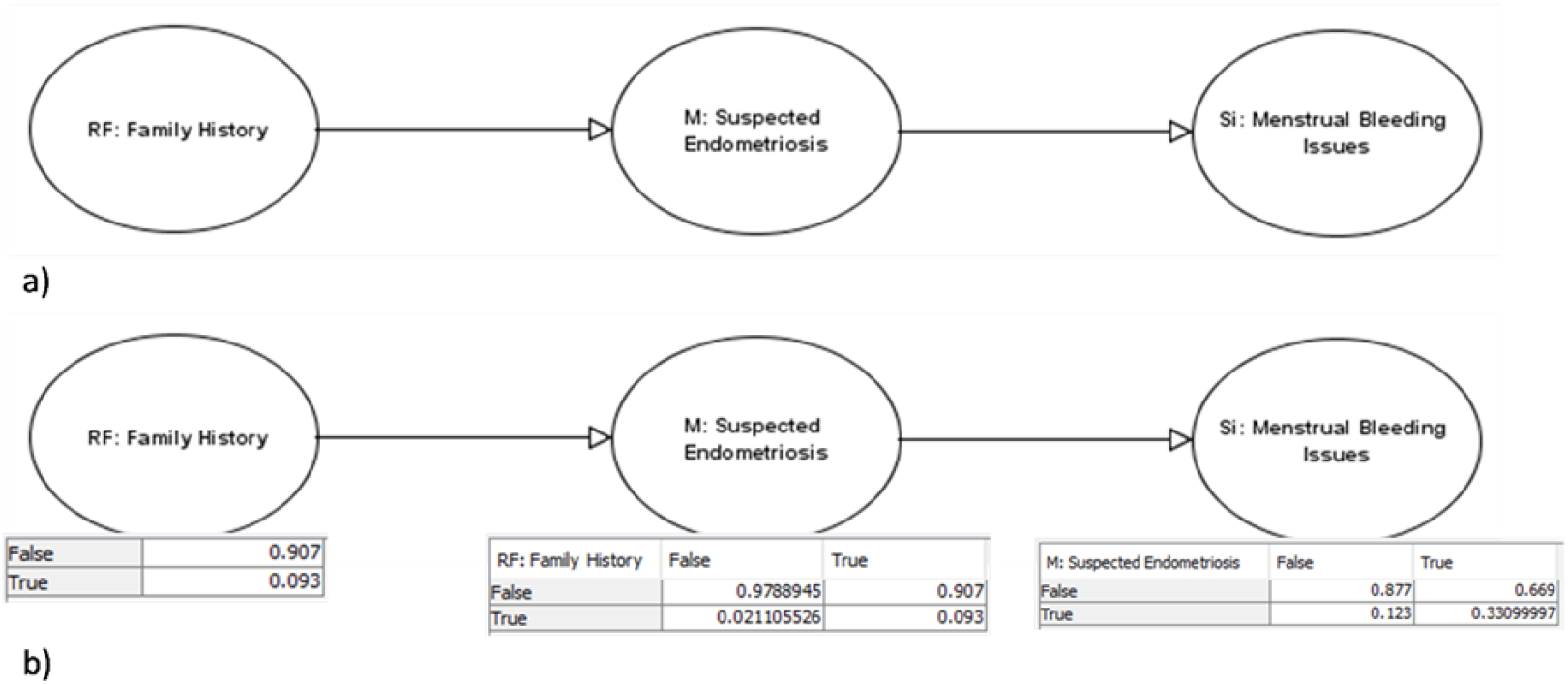
a) A three-node Bayesian Network casual model. b) A three-node Bayesian Network causal model with Node Probability tables displayed.

A benefit of using a Bayesian Network in medical applications is that once evidence is entered the unobserved variables in the model are updated. This is achieved by two reasoning methods: forward reasoning and backward reasoning. Forward reasoning follows the direction of the arc whereas backward reasoning is counter the direction. When the Bayesian Network structure represents the true causal relationships, rather than simple associations, they are referred to causal (prognostic) reasoning and diagnostic reasoning, respectively.

### Data and Selection of variables

A serious hurdle in practical applications of probabilistic methods is the effort required in model building and, in particular, of quantifying graphical models with numerical probabilities (Druzdzel & Díez, 2004). As reported medical symptoms can vary significantly between hospitals and general population, it is recommended that models are built using population studies. As defined by Holt & Weiss (2000), ‘definite endometriotic disease’ includes ovarian endometriomas, pelvic endometriotic lesions over 5mm deep and pelvic endometriotic lesions with adhesions not attributable to other causes. Studies without clinical confirmation of the condition were excluded. Where available, data were collected from various population-control studies including Burton et al.’s (2017) case-control study using primary care electronic health records to isolate early pointers towards a diagnosis of endometriosis, Bérubé et al.’s (1998) study of endometriotic feature in infertile women, and Trabert et al.’s (2011) study of diet and risk of endometriosis, where diagnosis of endometriosis had been diagnostically made by laparoscopic procedure. Data to inform about the impact of socio-economic on the access of health care were taken from Williams et al.’s (2010) study ‘Race, Socioeconomic Status and Health: Complexities, Ongoing Challenges and Research Opportunities.’ Data regarding the patient’s satisfaction with care received and the impact endometriosis has on their quality of life was taken from a study exploring a patient’s journey to an endometriosis diagnosis (Lamvu, et al., 2020).

### Medical Idioms

The method of following applicable and reusable medical reasoning patterns, proposed by Kyrimi et al., was used to produce the diagnostic Endometriosis BN causal model. The use of medical idioms has been identified as a method that helps bridge the barriers between medical knowledge and decision science as it simplifies the task of data gathering and construction of the Bayesian network (Kyrimi, et al., 2020).

It is important to use classifications that represent the information that clinicians would normally use for describing a condition, therefore there is benefit in using medical idioms, such as Condition (C), Manifestation (M), Risk factors (RF), Treatment (T), Co-morbidities (CC) and Complications (Cm), to construct the model as shown in Figure 2. For clinical decision-making, it is necessary for the medical BN to capture all relevant clinical variables and follow the causal mechanisms of the medical problem as reflected in the human reasoning process (Kyrimi, et al., 2020). The model was constructed using AgenaRisk software (www.agenarisk.com). The model is available for download at www.eecs.qmul.ac.uk/~norman/Models/ (filename “endo.cmpx2) and can be run in the free trial version of AgenaRisk

**Figure 2.**
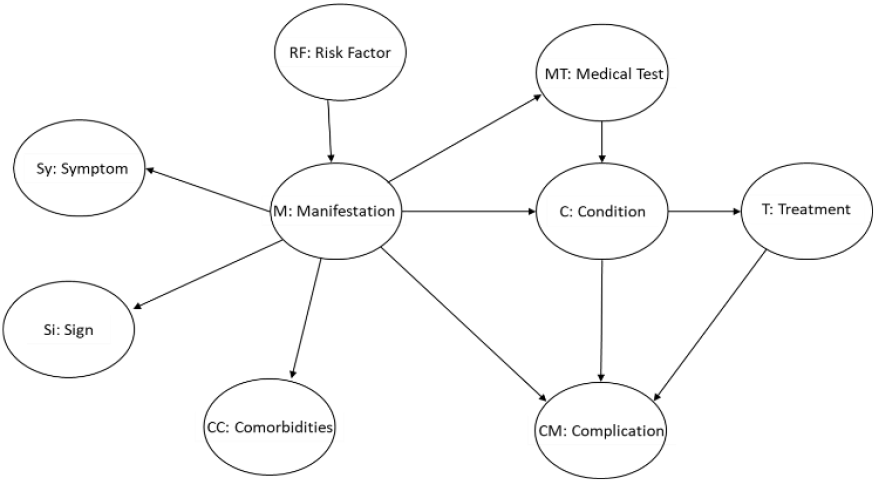
How medical idioms can be used to construct a Bayesian Network causal model.

### Implementation

Figure 3 displays the diagnostic Endometriosis BN causal model schematic. The starting point for building the model was to create the manifestation, ‘M: Suspected Endometriosis.’ The manifestation idiom models the uncertain causal relationship between the condition and the related manifestation variables, i.e. sign, symptoms and medical tests (Kyrimi, et al., 2020). The condition should exist before the effects are manifested; therefore, it was necessary to ascertain the marginal probability of suspected endometrioses from relevant risk factors.

**Figure 3.**
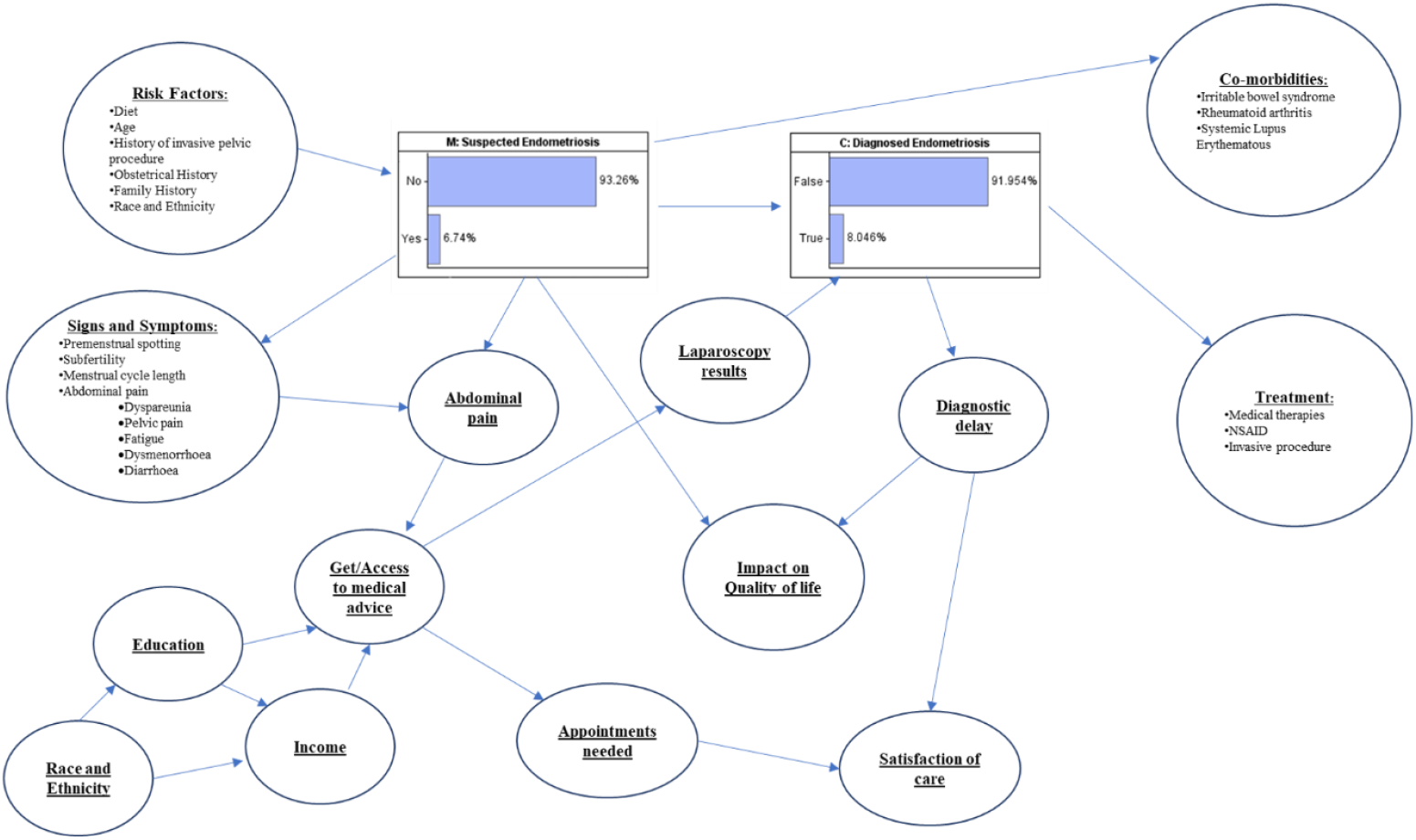
Endometriosis diagnosis model schematic.

Risk factors are part of a condition’s aetiology, as with many medical conditions, it is difficult to detect and correctly identify the underlying cause of endometriosis (Kvaskoff, et al., 2014). Risk factors are comprised of observable attributes, characteristics or exposures that increase a likelihood of developing a medical condition, or a manifestation of it. The risk factor idiom models the uncertain relationships between an observable risk factor and the variables it affects (Kyrimi, et al., 2020). Diet studies have weaknesses as the data collected are generally elicited from questionnaires and require patients to recall their eating habits; such a retrospective assessment by the patient can lead to measurement errors. Furthermore, different dietary patterns from different countries has led to contradictory results. Trabert et al.’s (2011) case-control study on diet and risk in endometriosis has been included in the endometriosis model due to its size and population-based design. This study found decreased endometriosis risk with increased intakes of total fat and dairy and increased risk associated with higher intake of β-carotene, a red-orange pigment found in plants and fruit such as carrots and sweet potato, and fruit (Trabert, et al., 2011). Bérubé et al.’s (1998) study of characteristics related to endometriosis in infertile women found the prevalence of minimal or mild endometriosis was higher in women aged 25 years or older. Women who had ‘gravida >0, para =0’ pregnancies, i.e. pregnancies that did not survive to a gestational age of 24 weeks, were more likely to have endometriosis (Bérubé, et al., 1998). Ashrafi et al.’s study evaluating the risk factors associated with endometriosis in infertile women confirms these risk factors and adds family history of endometriosis (OR: 4.9, CI: 2.1-11.3, P<0.0001) and history of pelvic surgery (OR: 1.9, CI: 1.3-.7, P<0.001) as potential risk factors (Ashrafi, et al., 2016). Figure 5 shows the risk factor nodes as parent nodes to the ‘*M: Suspected Endometriosis*’ node. In this instance ranked nodes were used to avoid having to manually define the Node Probability Table of ‘*M: Suspected Endometriosis’* cell by cell. Ranked nodes have order-preserving meaning, in this case ‘worst-to-best’ with regards to the level of risk. When a node is specified as such, no matter what the state labels or how many states a node has, there is an assumption that there is an underlying numerical scale that goes from 0 to 1 in equal intervals (Fenton & Neil, 2019). The ranked node ‘*M: Suspected Endometriosis’* is defined as a weighted sum, where the weight has been defined by the empirical evidence regarding each risk factor. To produce the NPT, a truncated Normal distribution is used, this is a flexible distribution that can accommodate a wide range of BN fragments involving a ranked node with ranked parents, as shown in Figure 4. The marginal probability of suspected endometriosis is 6.74%, reflecting its prevalence in the general population (Overton, et al., 2007). When all risk factors are true/highest this probability increases to 16.406%, as shown in Figure 10. A parent node *‘RF: All unknown causal factors’* is included with a high weighting compared to the other risk factors. This node will never be observed, it represents the confounding variables and at its maximum level would ensure ‘*M: Suspected Endometriosis’* is true.

**Figure 4.**
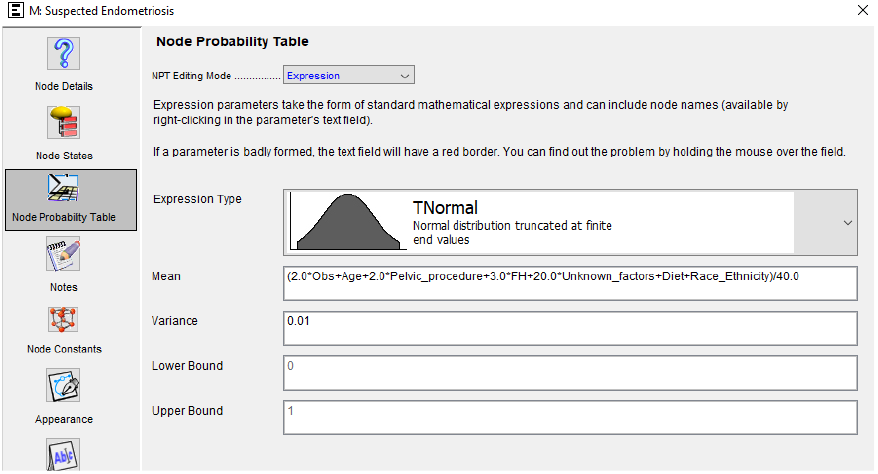
M:Suspected Endometriosis Node Probability table

**Figure 5.**
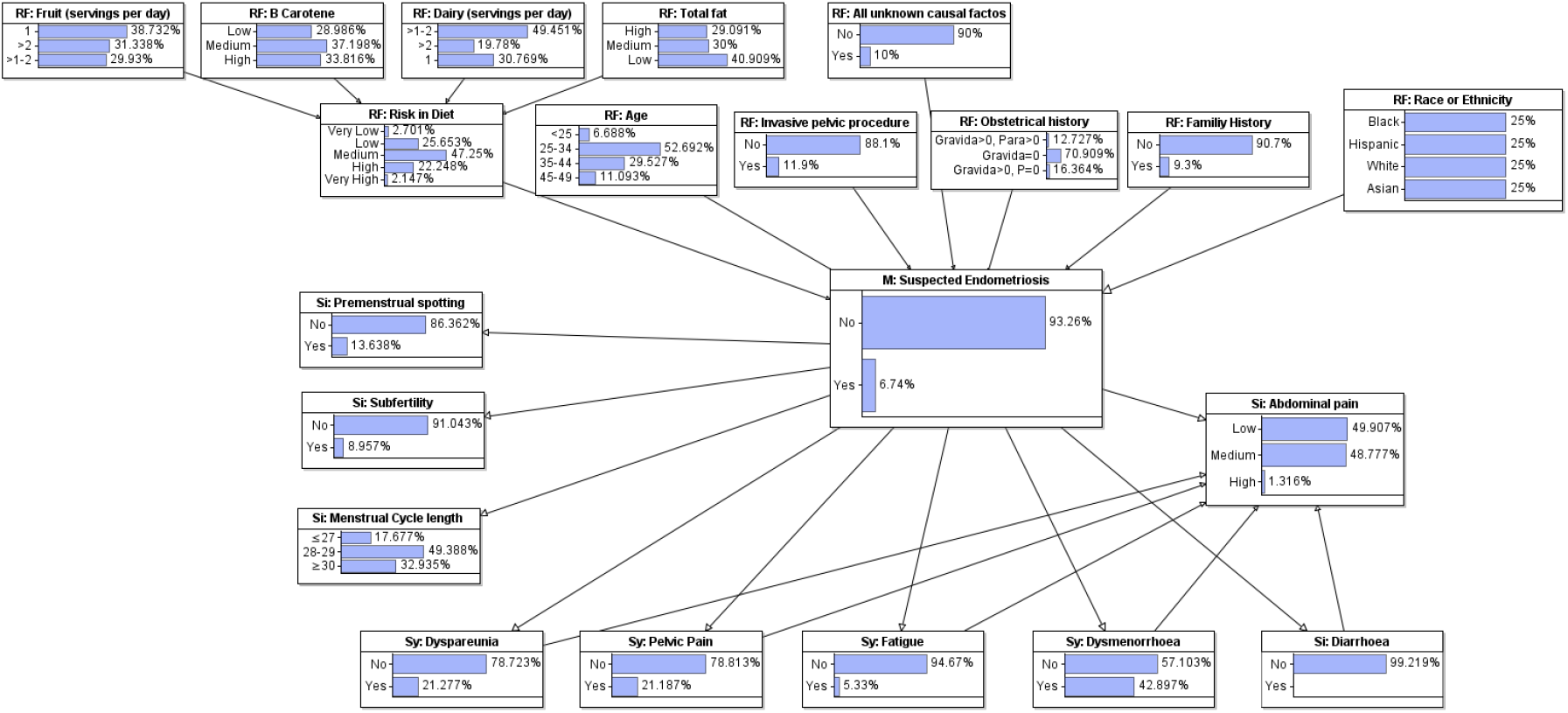
Diagnostic endometriosis model displaying risk factors, signs and symptoms associated with the condition.

**Figure 6.**
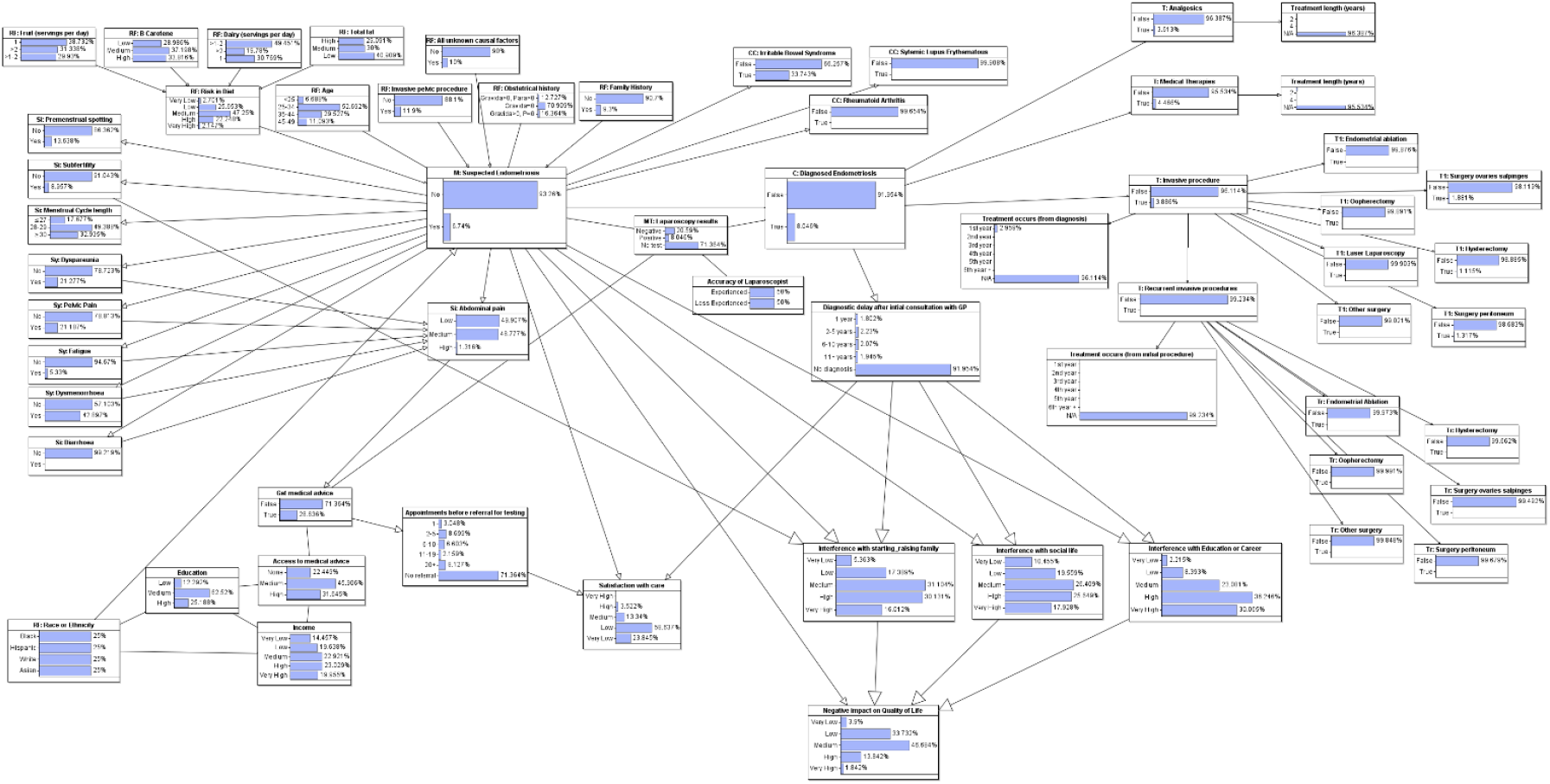
Full endometriosis diagnostic model.

**Figure 7.**
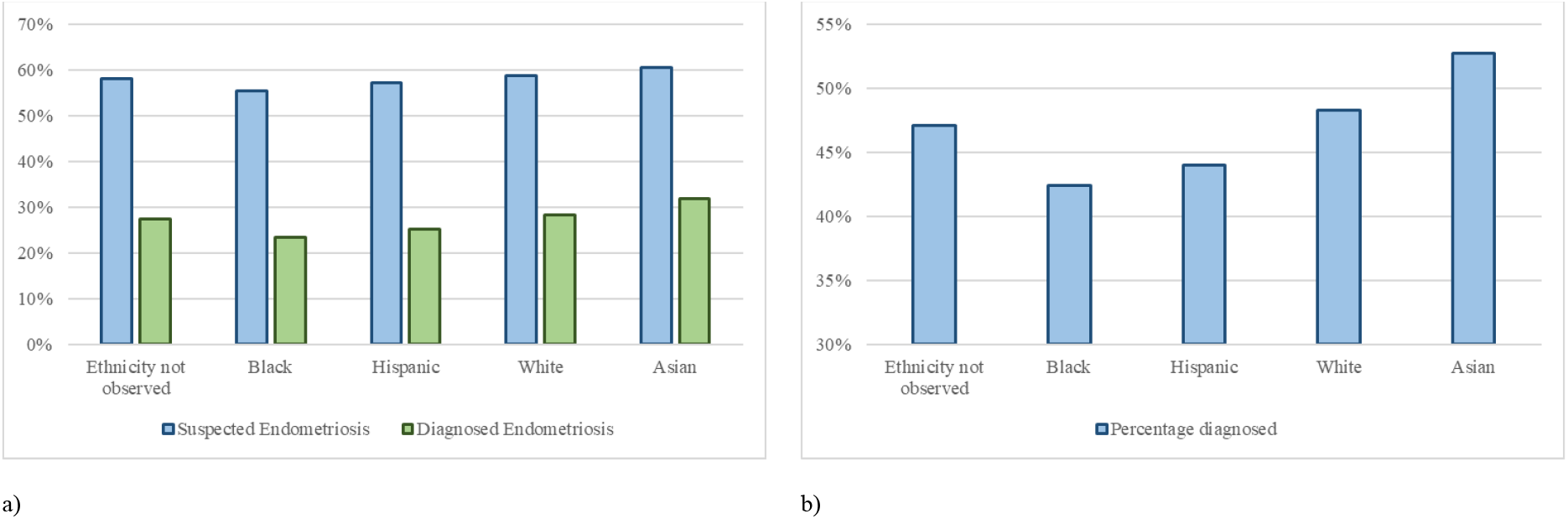
a) Suspected endometriosis probability compared with diagnosed endometriosis probability, grouped by race or ethnicity. b) Percentage of patients with endometriosis who get diagnosed, grouped by race or ethnicity.

**Figure 8.**
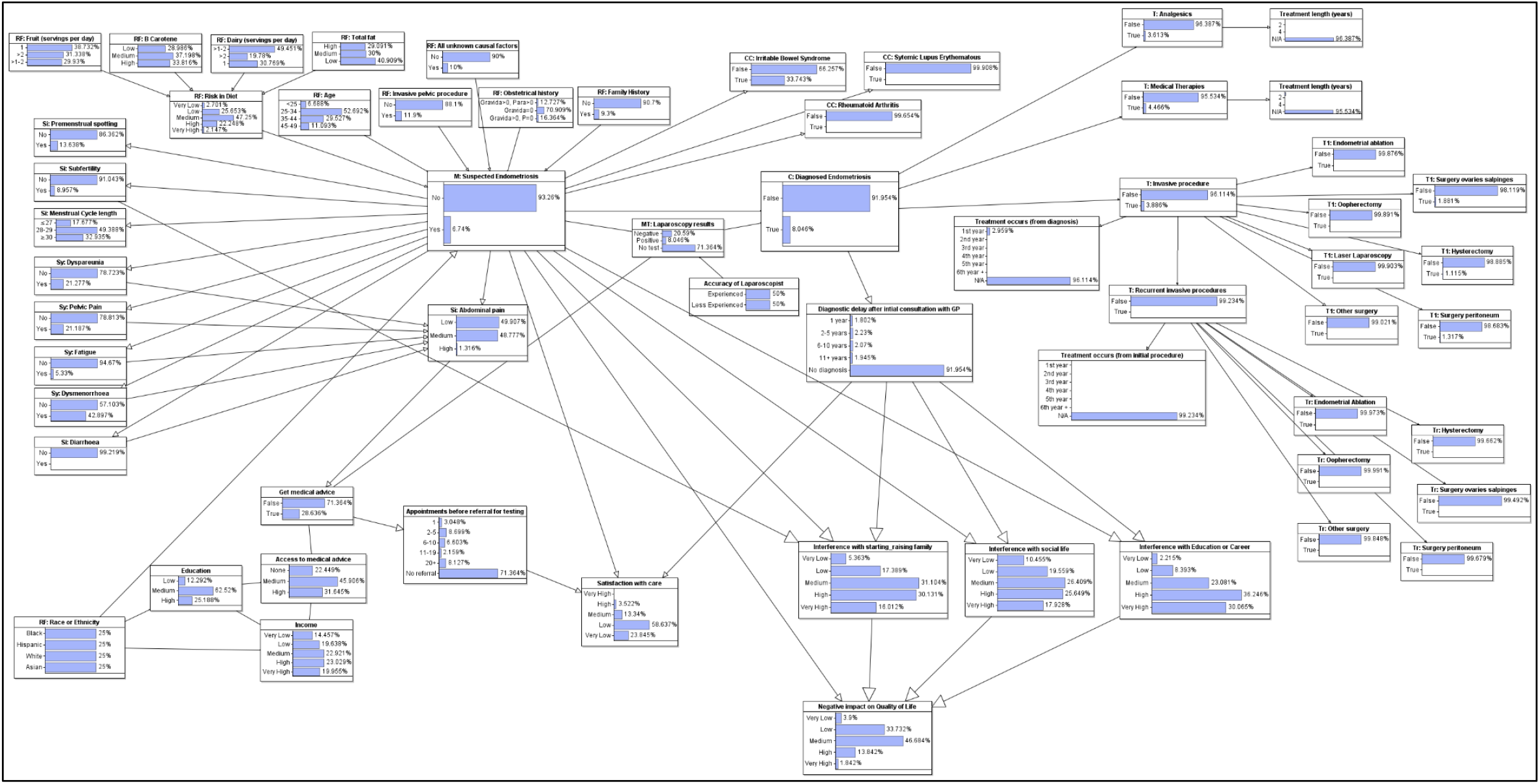
Full Endometriosis Diagnostic Bayesian Network model.

**Figure 9.**
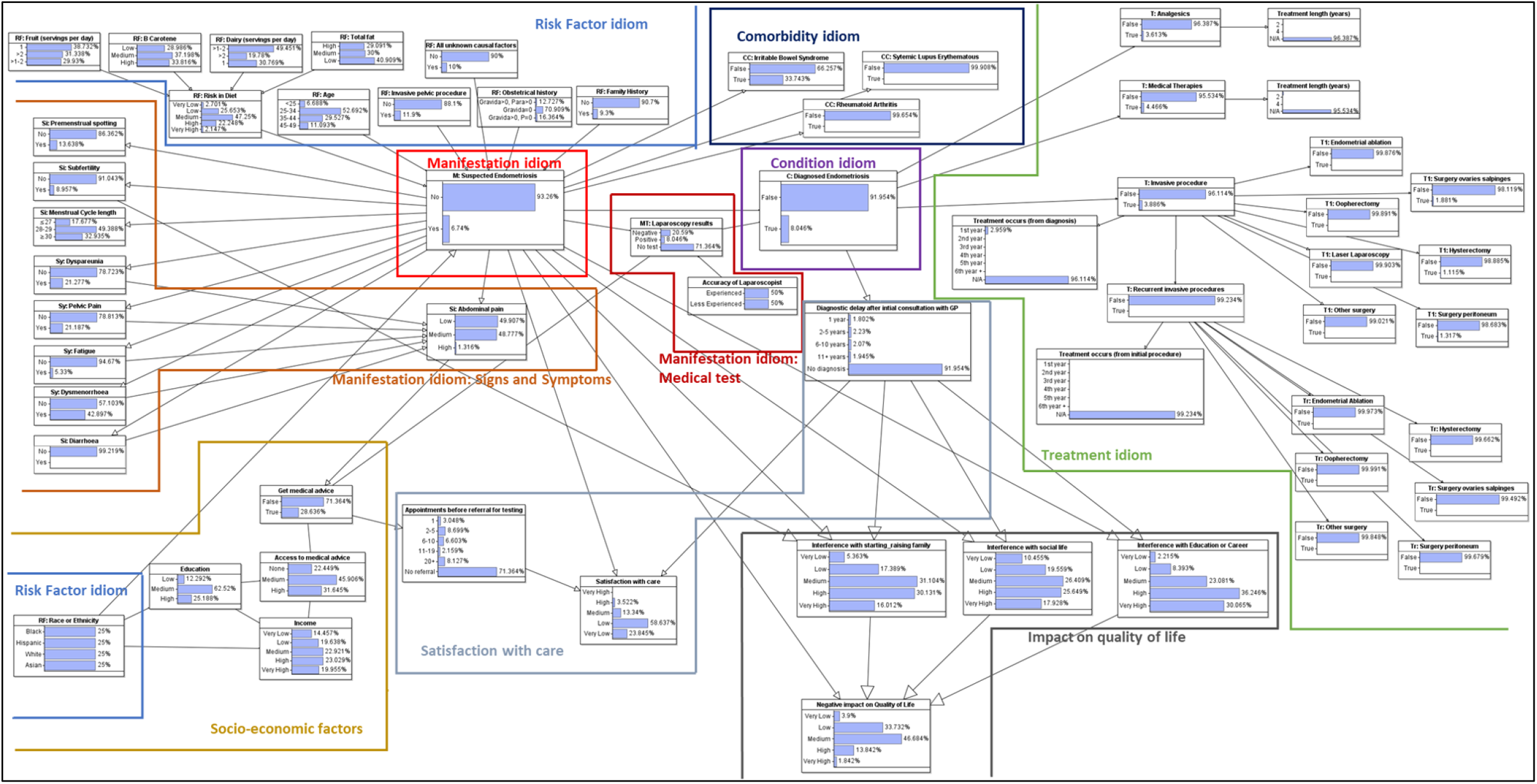
Full Endometriosis diagnostic model with medical idioms indicated.

**Figure 10.**
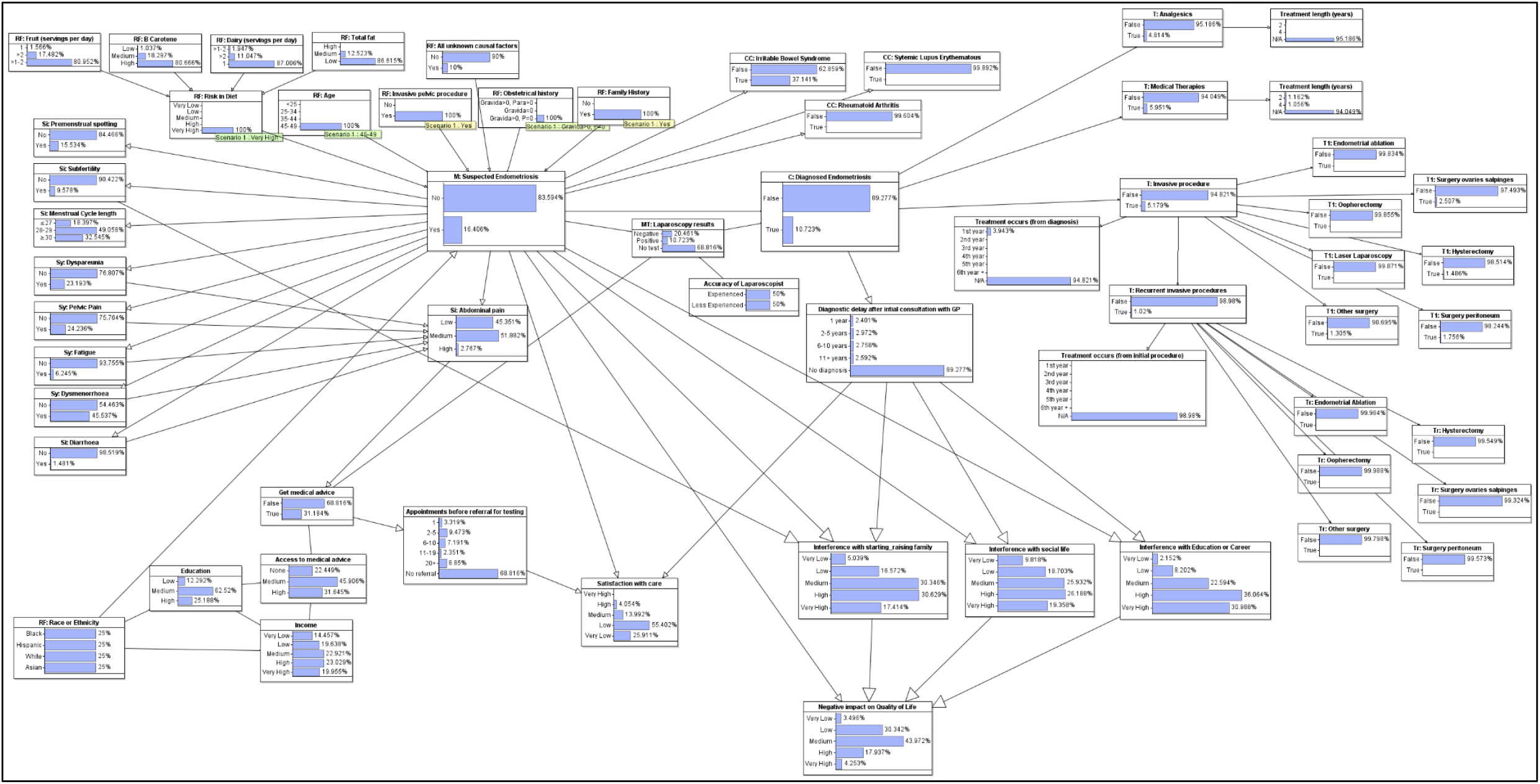
Endometriosis diagnostic model showing all risk factors observed.

To determine the condition in the Endometriosis model, ‘*M: Suspected Endometriosis*,’ the observable consequences of the condition were added; symptoms, signs, and medical tests. Figure 5 shows the presence of endometriosis manifests into premenstrual spotting, subfertility, dyspareunia, pelvic pain, fatigue, dysmenorrhoea, and diarrhoea and menstrual cycle length. Symptoms (Sy) are subjective feelings which are apparent only to the patient and cannot be measured or quantified by the clinician. Pain and fatigue are symptoms that require self-reporting from the patient. In contrast, signs (Si) are objective indications that are observable to the clinician. The sign and symptom nodes are, again, ranked nodes that are children to ‘*M: Suspected Endometriosis*.’ Throughout most studies analysing features of endometriosis, abdominal and pelvic pain was identified as the principle symptom of the disease (Burton, et al., 2017; Grundström, et al., 2016; Rolla, 2019). The ‘*Sy: Abdominal pain’* node is a ranked node describing pain as ‘low’, ‘medium,’ or ‘high.’ It is the child of ‘*M: Suspected Endometriosis*’ and symptoms relating to pain. It is the parent of ‘*Get medical advice’*, the greater the level of pain the more likely the patient is to seek medical care, be referred for a test and therefore be diagnosed with endometriosis.

The condition in the Endometriosis model is the diagnosed ailment of endometriosis; ‘*C: Diagnosed Endometriosis*.’ It is a Boolean node, stating true or false, the probability of a patient being diagnosed with endometriosis. It is the child node of ‘*M: Suspected Endometriosis’* and *‘MT: Laparoscopy results*.’ Diagnosis can be true only with laparoscopy results confirming endometriosis, with or without suspected endometriosis being true. It is the parent node to the treatment nodes as treatment can only commence when diagnosis is made.

The medical test (MT) idiom indicates the procedure performed to diagnose the medical condition (Kyrimi, et al., 2020). Diagnosis of endometriosis is made following a laparoscopic inspection of the pelvis. It is considered the gold standard for confirmatory diagnosis, with histologic conformation after biopsy (Rolla, 2019; Parasar, et al., 2017). A study by Wykes et al. (2004) endeavoured to provide precise diagnostic accuracy estimates. It was concluded that the overall sensitivity was 94% and specificity was 79% for the accuracy of diagnosis by laparoscopy. This data is input in a labelled node ‘*MT: Laparoscopy results*,’ a child node of ‘*M: Suspected Endometriosis*,’ and *‘Get medical advice*,’ and a parent node to ‘*C: Diagnosed Endometriosis*.*’* The inaccuracy of each manifestation variable is accounted for as a false positive and false negative rate. However, the reliability of the patient reporting their symptom, or the accuracy of the clinician performing tests, can be another source of inaccuracy (Kyrimi, et al., 2020). It was important, therefore, to introduce an accuracy node, a manifestation reliability idiom, for the experience of the laparoscopist conducting the examination. It was reported in 2002 that experienced laparoscopists have an 86% accuracy and less experienced laparoscopists have a 41% accuracy when compared to histologic biopsy results (Wood, et al., 2002). A labelled node was used ‘*Accuracy of Laparoscopist’* to indicate the level of experience and when the node probability table of ‘*MT: Laparoscopy results’* was updated to reflect this it revealed a 63.5% probability that a patient will be diagnosed with endometriosis.

The treatment (T) idiom represents the clinical management to treat the condition (Kyrimi, et al., 2020). In their first year following diagnosis of endometriosis, 55.5% of patients are prescribed medical therapies (Cea Soriano, et al., 2017). The most prescribed medication were combined oral contraceptives (COCs). Medical therapies also includes progestogen-only pills (POPs), implants, and injections, gonadotropin-releasing hormone (GnRH), copper and non-specific intrauterine devices (IUDs) among others. Pain medication was prescribed to 44.9 % of patients (Cea Soriano, et al., 2017). Non-steroidal anti-inflammatory drugs (NSAIDs), to treat pain, were also commonly prescribed. This list includes codeine, tramadol, and opioids. 48.3% of patients with confirmed endometriosis require invasive treatment, with most of this treatment taking place in the first three years following diagnosis. The most common surgical treatment is surgery to the ovaries and/or salpinges, also referred to as fallopian tubes. 19.7% of patients require further invasive treatment following an initial procedure (Cea Soriano, et al., 2017). 28.7% of patients underwent hysterectomy during their initial invasive procedure, 44.2% underwent hysterectomy following recurrent invasive procedures (Cea Soriano, et al., 2017). The treatment nodes can be identified by ‘T’ before the node name, for example ‘*T: Medical Therapy*.’ These nodes are binary and are child nodes from the parent node ‘*C: Diagnosed Endometriosis*.’ Recurrent treatment, invasive procedures that has occurred after an initial invasive procedure has already been performed, has been given the indicator ‘Tr’, for example ‘*Tr: Hysterectomy*.’

The race and ethnicity of the patient has long been touted as a risk factor for endometriosis with black women less likely to be diagnosed with endometriosis and Asian women more likely to be diagnosed when compared to white women. However, in women presenting with infertility, there was no significant difference in endometriosis prevalence between white and black women (Bougie, et al., 2019). It seems likely that race and ethnicity may influence the ability to access healthcare and obtain appropriate management for endometriosis through a combination of socio-economic issues (Williams, et al., 2010). Regardless, this causes an implicit and explicit bias among the medical community, which can influence a patient’s chance of a timely and accurate diagnosis of endometriosis, depending on their ethnicity. Statistical data was taken from the United States Census Bureau (2009) to create ranked nodes displaying education, income, and access to medical to generate a node to indicate whether a patient would be able to access medical care regardless of the probability of suspected endometriosis, Figure 3.

To determine how satisfied patients are with the care they get on their diagnostic journey a ranked node with a weighted mean was created, ‘*Satisfaction with care*.’ It is the child node of ‘*Appointments before referral for testing’*, ‘*M: Suspected Endometriosis*,’ and ‘*Diagnostic delay after initial consultation with GP*.’

Finally, nodes representing impact on area of life important to the patient were created. *‘Interference with starting_raising a family*’; a ranked node whose parents are ‘*RF: Subfertility*,’ ‘*M: Suspected Endometriosis*,’ and ‘*Diagnostic delay*’; and ranked nodes ‘*Interference with Education or Career*’ and ‘*Interference with social life*’; whose parents are both ‘*M: Suspected Endometriosis*’ and ‘*Diagnostic delay*.’ Finally, these interference nodes alongside ‘*M: Suspected Endometriosis’* are parents of *‘Negative impact on Quality of Life’* to determine how patients with endometriosis are affected in their day-to-day living. The information was pulled from a study exploring a patient’s journey to an endometrioses diagnosis (Lamvu, et al., 2020).

## Results

The final diagnostic BN causal model for endometriosis can be seen below in Figure 6, a larger image is available in *Appendix A* Figure 8. The primary objective of the model is to calculate the probability of having suspected endometriosis by observing risk factors, signs, and symptoms of the condition. The model then calculates the probability of the patient being surgically diagnosed with the condition. The secondary objective of the model is to predict the satisfaction of the patient from the care that they receive, and the impact endometriosis has on their quality of life. Scenarios and their results have been outlined below.

### Diagnosis and Treatment results

Images of the model displaying diagnosis results can be viewed in *Appendix B*. Prior to any observed risk factors, the risk of suspected endometriosis is 7%, as seen in Figure 6, reflecting the prevalence of endometrioses is the general population. When risk factors including diet, age, previous invasive pelvic procedure, obstetrical history, and family history are observed the risk of endometriosis rises to 16%, as shown in Figure 10. A node representing all unknown causal factors has been included. This node will never be observed, but if observed to be true, will ensure the probability of endometriosis is true.

When all signs and symptoms are observed the probability of endometriosis is not in doubt, the probability produced is 99.69%, the level of abdominal pain is high at 80% and the likelihood of the patient seeking medical advice rises to nearly 70%, as shown in Figure 12. The probability of being diagnosed with endometriosis is dependent on being referred for a laparoscopic scope and the accuracy of the laparoscopist. Figure 12 shows that without observations the probability of being diagnosed with endometriosis is 44%. If ‘get medical advice’ is observed to be true, the probability of being diagnosed increases to 63%, as shown in Figure 13. If an experienced laparoscopist is observed this figure rises to 86%, an unexperienced laparoscopist observed drops the figure to 41%, Figure 14.

**Figure 11.**
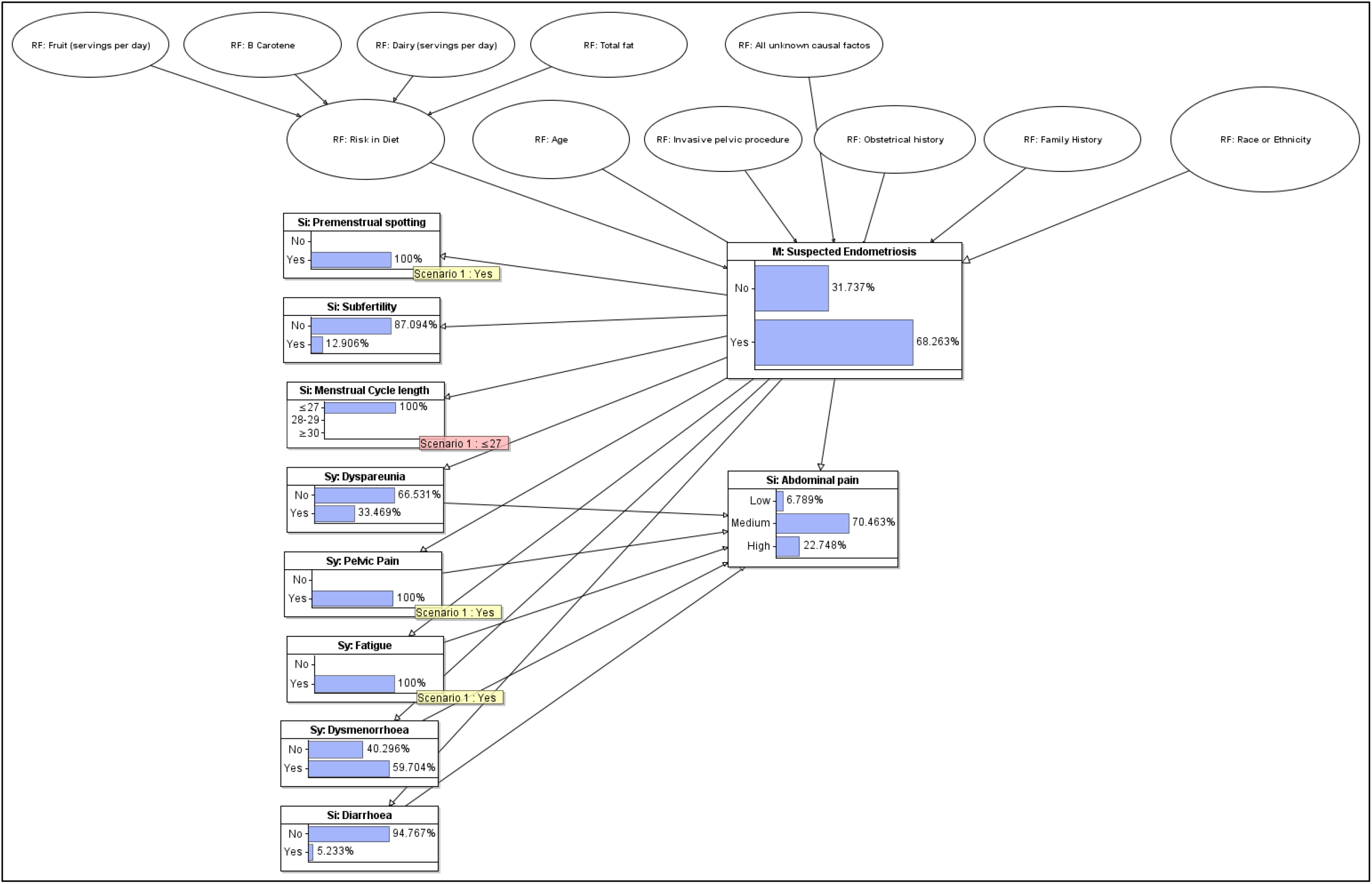
Section of Endometriosis diagnostic model showing how selecting a few symptoms can lead to M: Suspected Endometriosis to be largely true.

**Figure 12.**
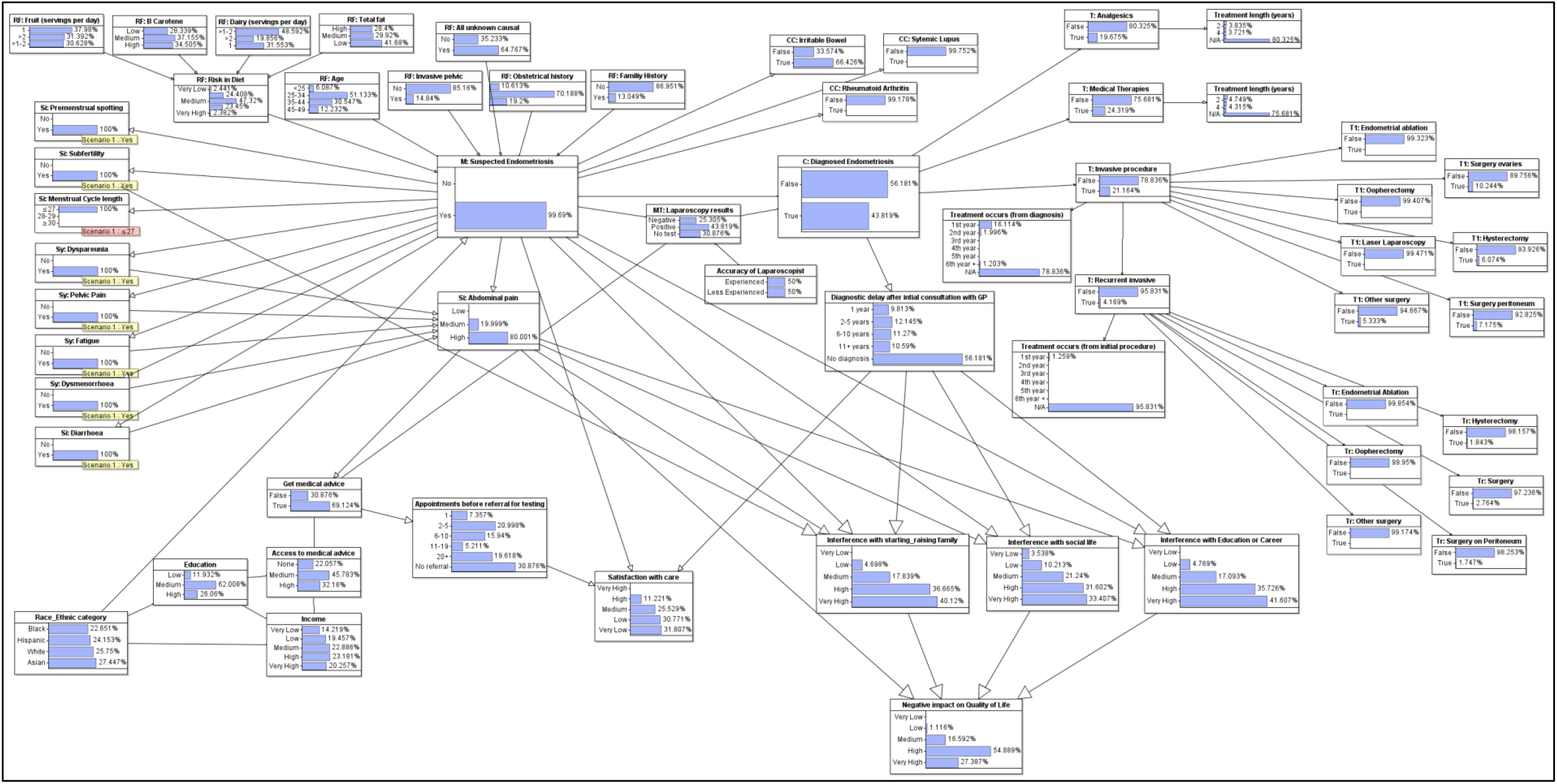
Endometriosis diagnostic model with all signs and symptoms observed.

**Figure 13.**
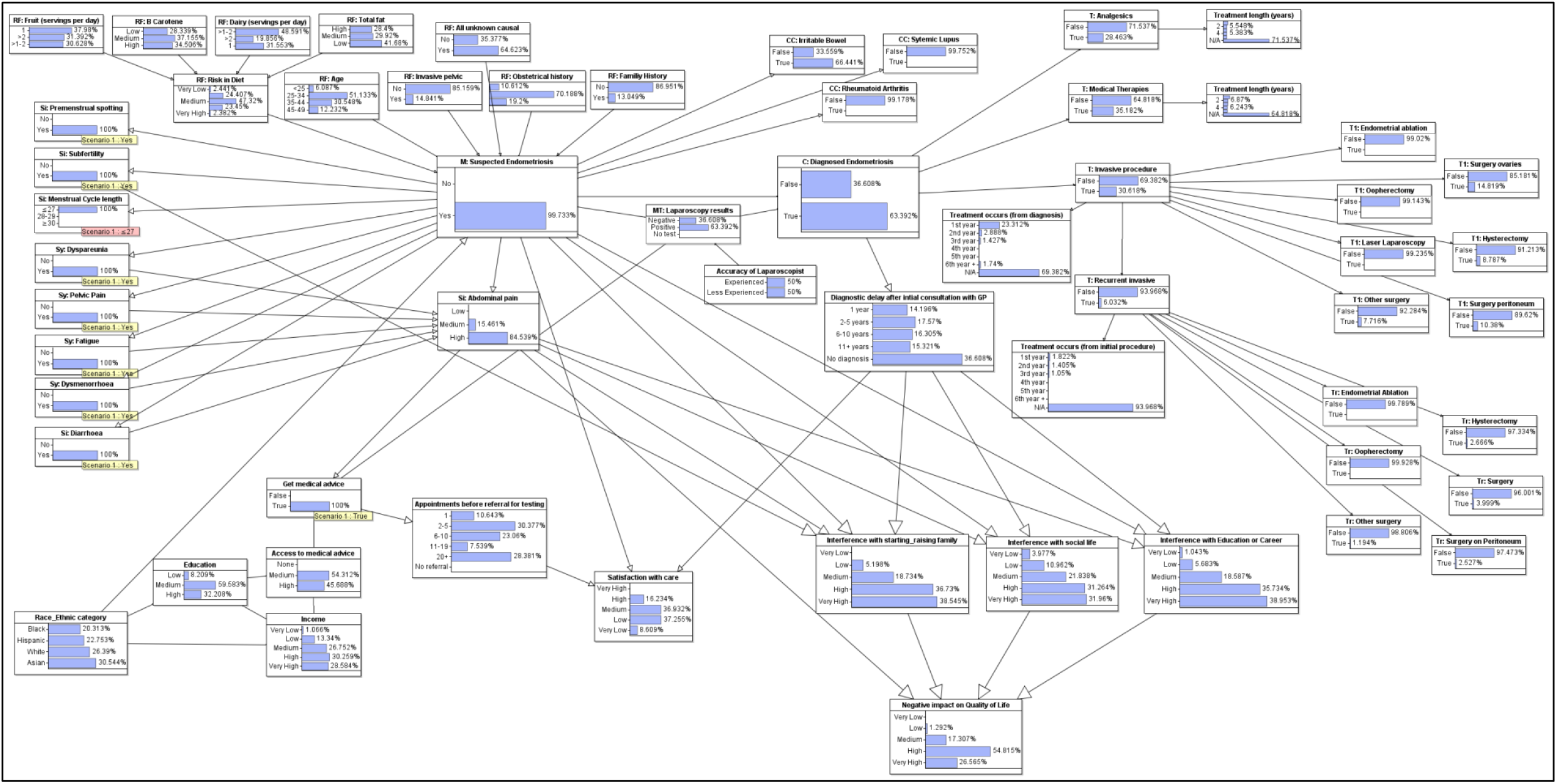
Endometriosis diagnostic model with ‘Get medical advice’ observed.

**Figure 14.**
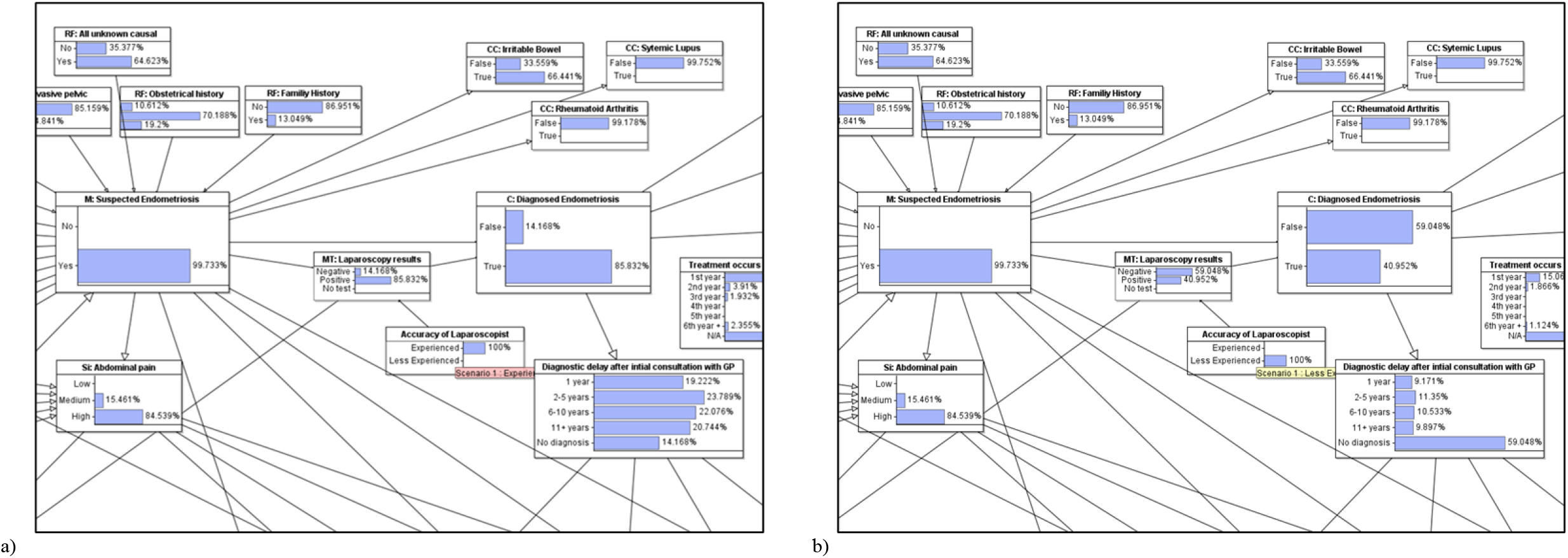
*a)* Endometriosis diagnostic model with experience laparoscopist observed. b) Endometriosis diagnostic model with less experienced laparoscopist observed.

Once diagnosis is observed to be true the most likely treatment is medical therapies, most often combined oral contraception, at 56%, Figure 15. Analgesic treatment lies at 45%, and invasive procedure probability is 48%. By observing the treatment categories, medical therapy and analgesic treatment regularly take place over 5 years. The most likely invasive procedure a patient will undergo is surgery to the ovaries and salpinges, at 48%. This is followed by peritoneum surgery, the lining of the abdominal cavity, at 34% and hysterectomy, the removal of the uterus, at 29%. It is important to note that there is a 20% chance that the patient will have to undergo recurrent invasive procedures Figure 16. When recurrent procedure is observed, the probability of surgery to the ovaries and salpinges is again the highest at 66%. This is followed by hysterectomy, at 44%, indicating that the delay may be due to factors such as wishing to produce children.

**Figure 15.**
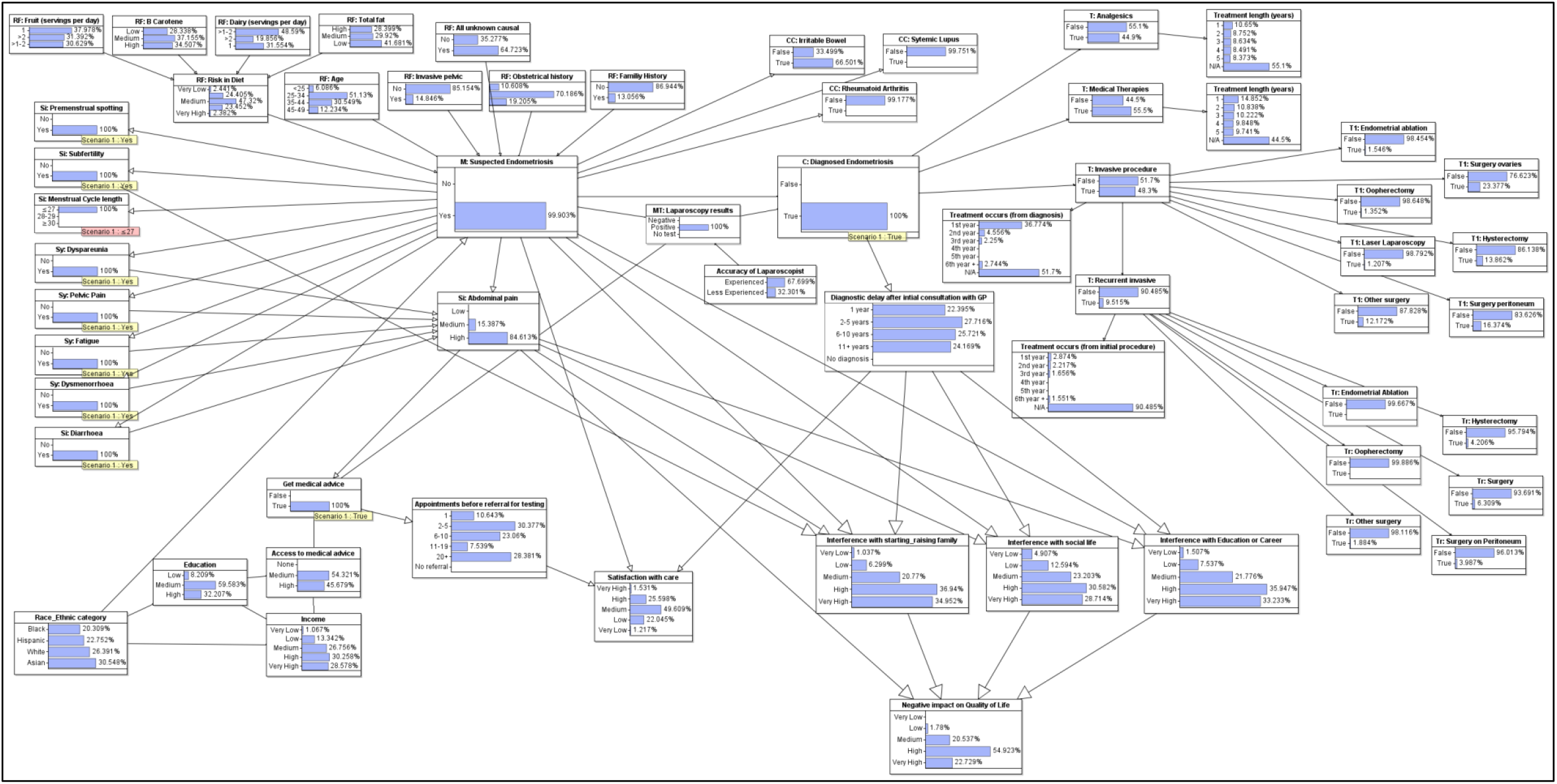
Endometriosis diagnostic model with ‘Diagnosed Endometriosis’ observed as true.

**Figure 16.**
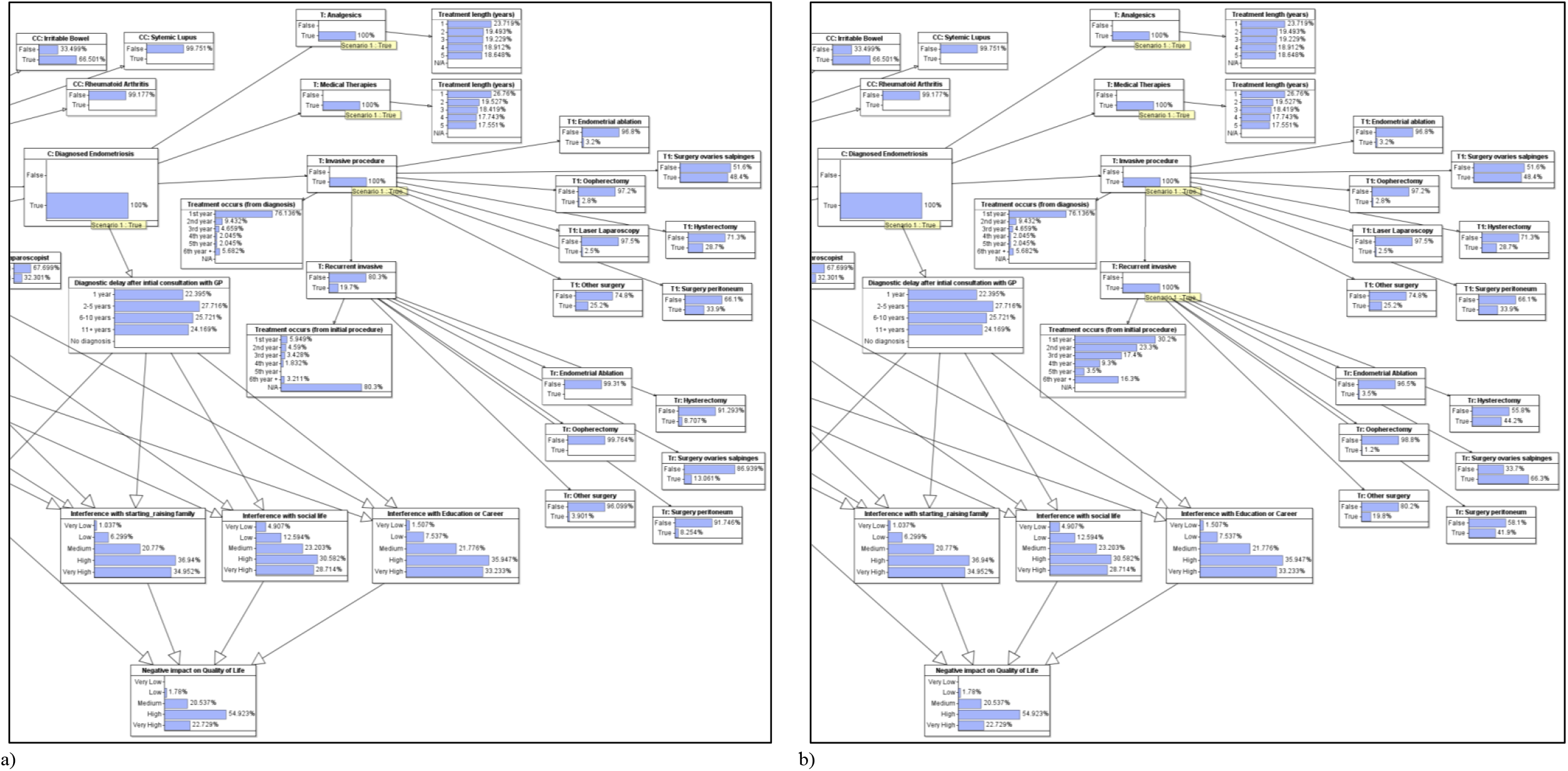
a) Endometriosis diagnostic model with treatments observed. b) Endometriosis diagnostic model with Recurrent invasive procedures observed

### Race and Ethnicity factors

The impact of race and ethnicity to the risk and potential diagnosis of endometriosis can be viewed in figures attached in *Appendix C*. Figure 17 a) shows that the probability of suspected endometriosis is 55% when ‘Black’ has been observed in the ‘*RF: Race or Ethnicity’* node. This figure rises to 57% when ‘Hispanic’ is selected, 59% when ‘White’ is selected, and 60% when ‘Asian’ is selected, as shown in Figure 17 b) and Figure 18, a) and b). This rise in risk reflects the influence of race or ethnicity on the prevalence of endometriosis, as discussed in literature above. Yet, the probability of receiving a confirmed diagnosis of endometriosis is also affected by race and ethnicity. The probability of confirmed endometriosis is 23% when ‘Black’ is observed, 25% when ‘Hispanic’ is observed, 28% when ‘White’ is observed and 32% when ‘Asian’ is observed. Figure 7 a) displays these figures on a bar chart. At first glance, the figures displayed on the chart look proportional to each other. However, when the percentage of patients with suspected endometriosis who are successfully diagnosed with endometriosis is calculated it becomes evident that patients with a Black and Hispanic background are less likely to get a successful diagnosis. Black patients are successfully diagnosed 42% of the time compared to Asian patients, who have a 53% probability of being diagnosed. This result shows that education, income, and access to healthcare have a direct influence on whether a patient will be diagnosed with endometriosis.

**Figure 17.**
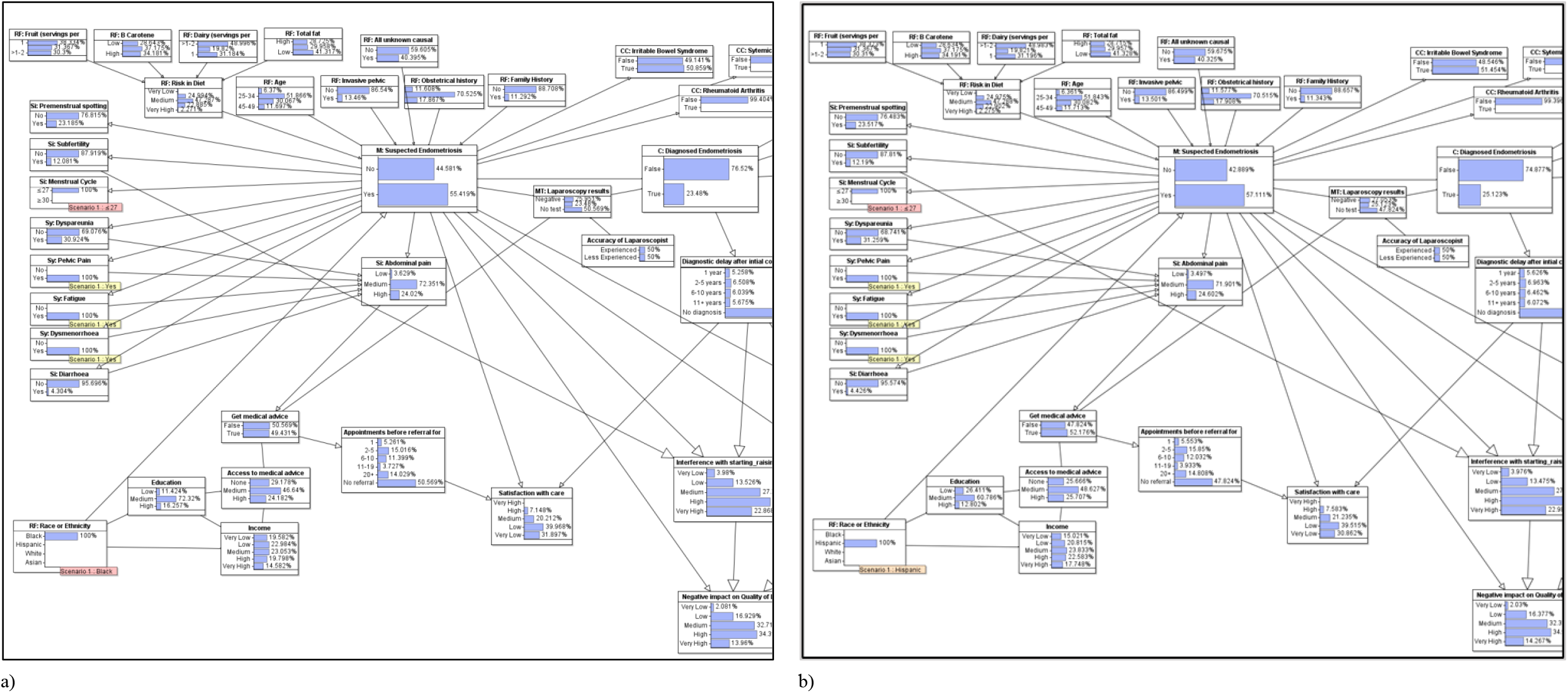
a) Suspected endometriosis probability 55%, Diagnosed endometriosis probability 23% when ‘Black’ observed in Race or Ethnicity node. b) Suspected endometriosis probability 57%, Diagnosed endometriosis probability 25% when ‘Hispanic’ observed in Race or Ethnicity node

**Figure 18.**
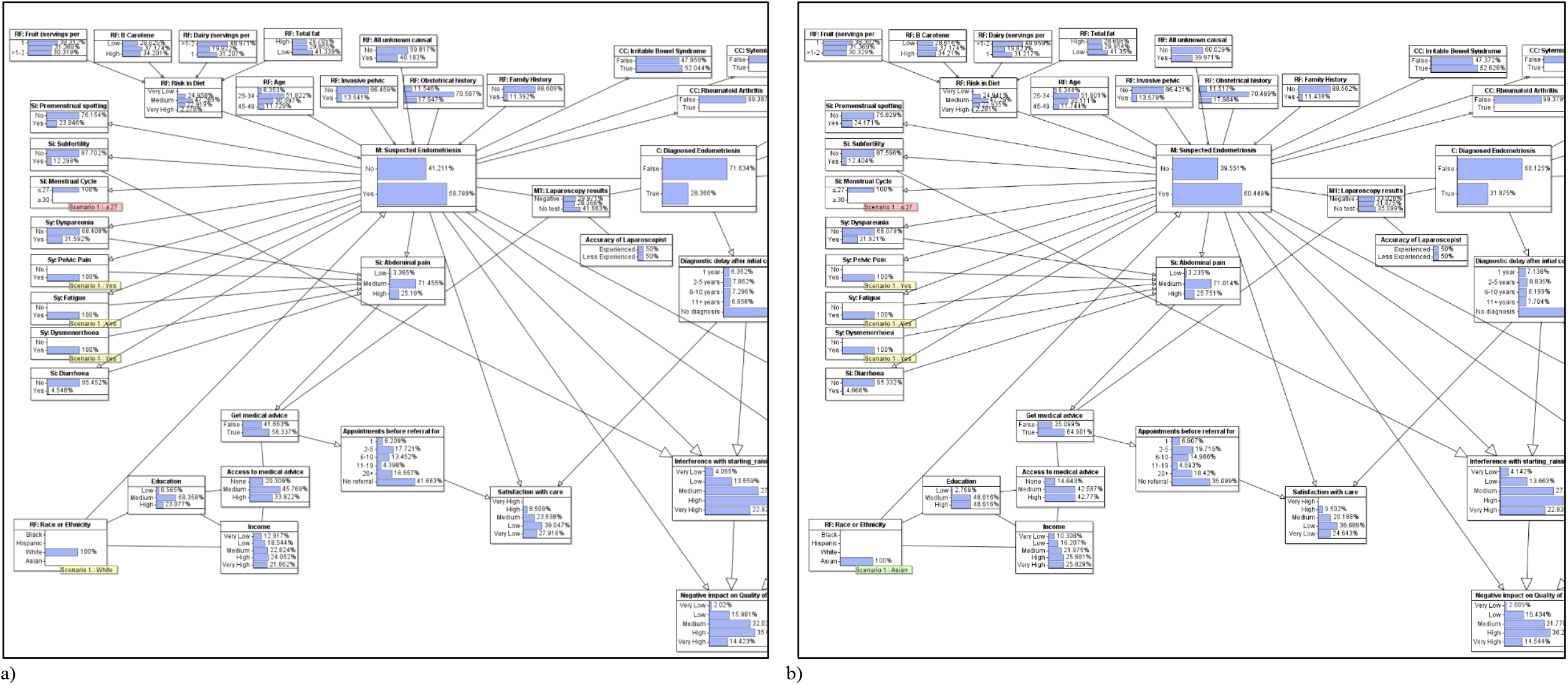
a) Suspected endometriosis probability 59%, Diagnosed endometriosis probability 28% when ‘White’ observed in Race or Ethnicity node. b) Suspected endometriosis probability 60%, Diagnosed endometriosis probability 32% when ‘Asian’ observed in Race or Ethnicity node

### Satisfaction with care

Please refer to *Appendix D* for figures relating to this scenario. When a patient does get medical advice there is a 30% probability that they will have to attend two to five appointments before being referred for testing, Figure 13. The second highest outcome from this observation is over 20 appointments before referral (28%). This is delay is likely due to the physician treating the symptoms rather than the condition and leads to frustration and lack of satisfaction with care for the patient (Lamvu, et al., 2020). When a patient is diagnosed with endometriosis there is a 28% probability that it occur two to five years following their initial consultation with a general practitioner. There is a 24% probability a patient will have to wait eleven or more years for diagnosis. Satisfaction with care falls the longer a patient must wait for referral or diagnosis, there is a 59% probability that a patient will have a high level of satisfaction when diagnosed in two to five years. This drops to an 81% probability of low level of satisfaction when diagnosis is delayed to eleven years or more, as shown in Figure 19 and Figure 20. Figure 20 also shows the negative impact on the quality of life coming from having endometriosis, the level of pain experienced and its impact on starting and/or raising a family, the impact on learning and/or the patients career, and its impact on the patient’s social life.

**Figure 19.**
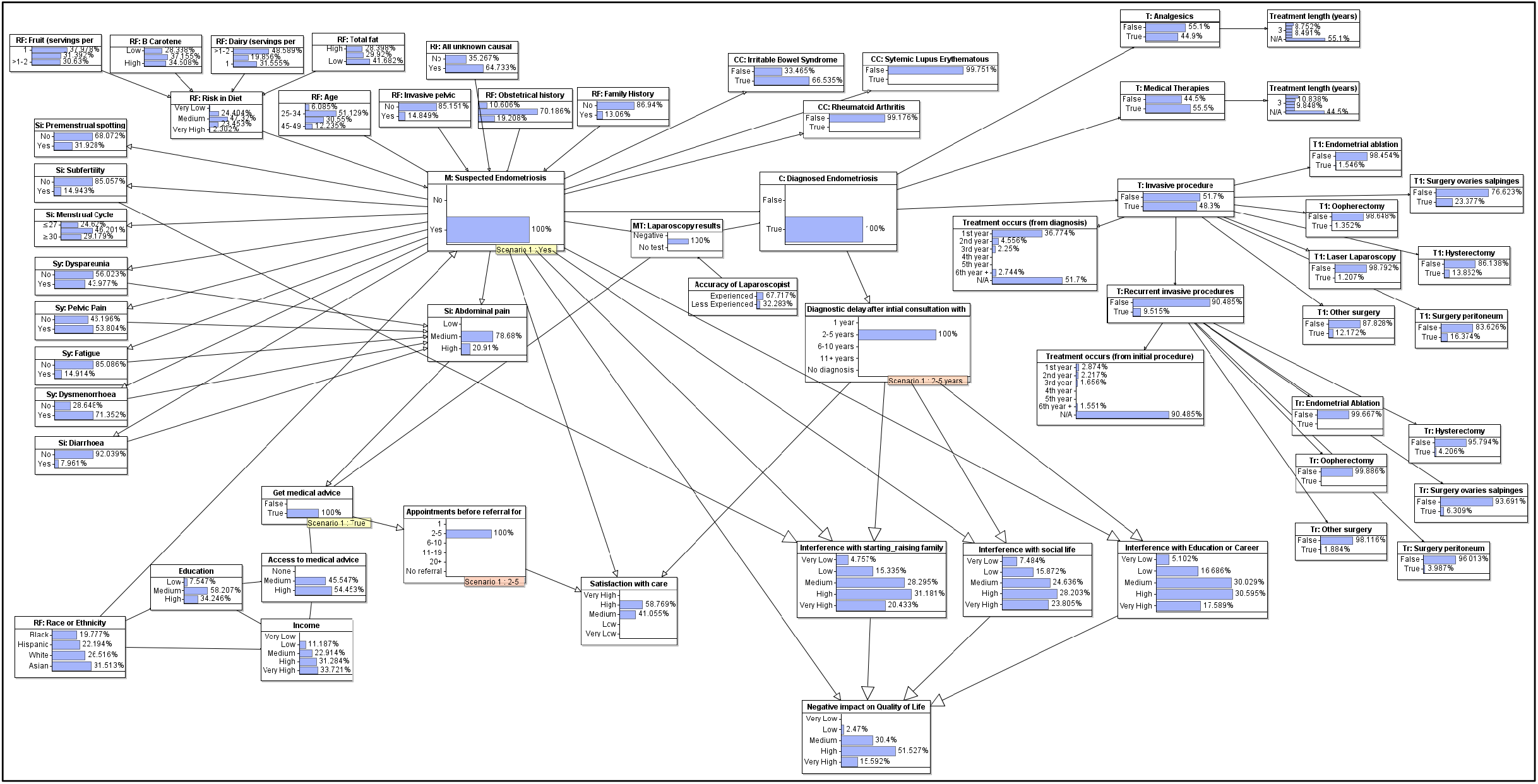
Endometriosis diagnostic model with ‘2-5’ observed for ‘Appointments before referral’ and ‘2-5 years’ for ‘Diagnostic delay’.

**Figure 20.**
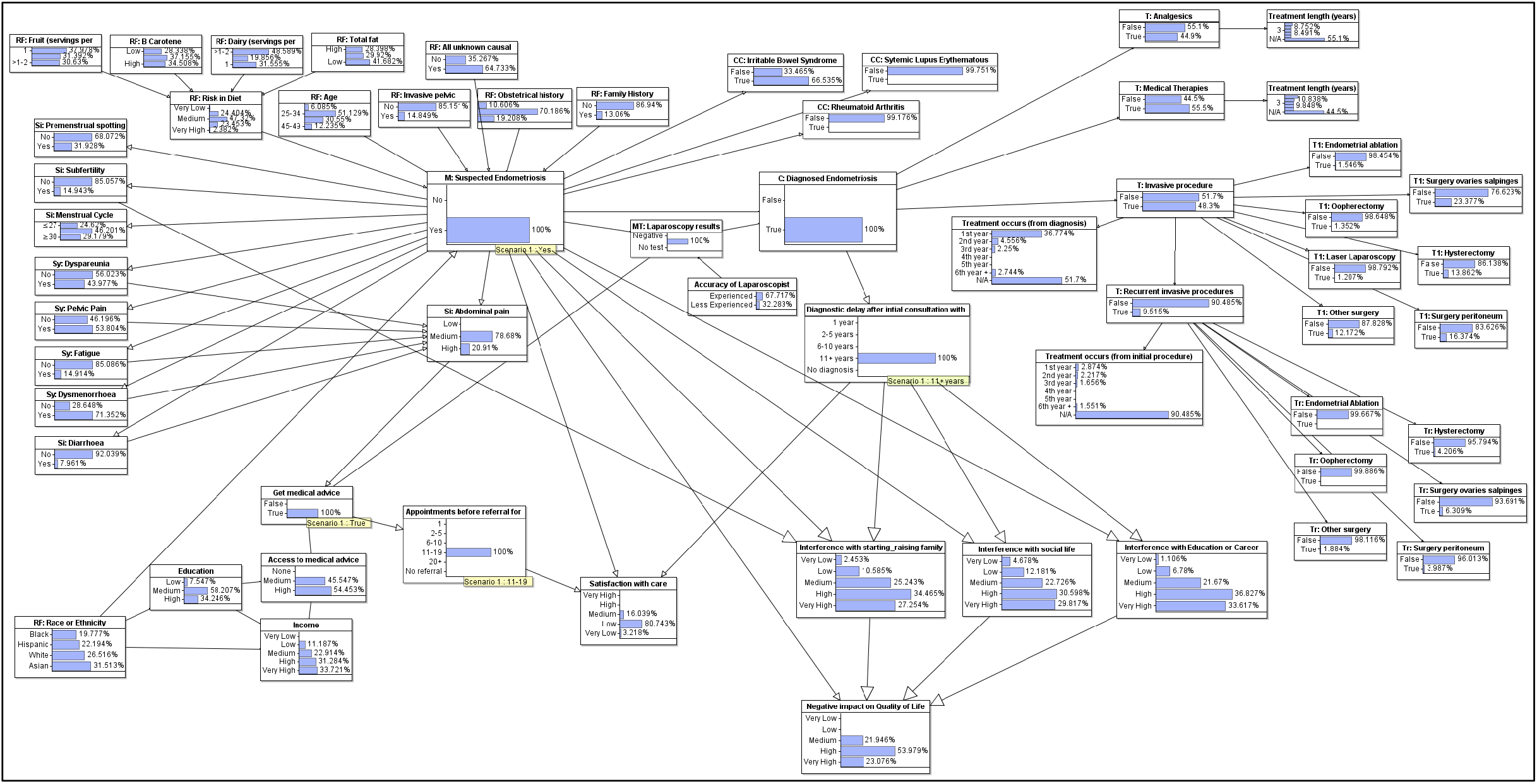
Endometriosis diagnostic model with ‘11-19’ observed for ‘Appointments before referral’ and ‘11+ years’ for ‘Diagnostic delay’.

## Discussion/Conclusion

In this study, it was illustrated how a Bayesian Network causal model can be used to aid clinical decision-making for the diagnosis of endometriosis by observing signs and symptoms of the condition. Due to its graphical nature, the interactions between the different variables built into the network are directly visualised and follow clinical reasoning through the use of medical idioms. A diagnostic model was achieved with supplementary analysis of socio-economic factors that may help or hinder possible diagnosis. Insight has also been provided into the impact endometriosis has on the patient’s quality of life, the impact on education and career, social life, and family life.

One limitation of the model is that, while it provides predictions of most likely treatment (given risk factors, signs, and symptoms), these predictions cannot be assumed to be ‘optimal’. Rather, it reflects past and current medical practice (much of which may be potentially sub-optimal or even flawed). To determine optimal treatment – taking account for example, of costs, side-effects, and the desire of the patient to maintain their fertility – the model needs to be extended to an influence diagram (Yet, et al., 2018).

It is also important to note that observing just a few symptoms means that ‘*M: Suspected Endometriosis’* is almost certainly true. We believe this may be because the data collected was tainted by confirmation bias. This may also expose a problem with the way endometriosis is currently being diagnosed, i.e. the condition is being defined by the symptoms. Yet, it is important to note that many women with endometriosis are asymptomatic, may not seek medical care or may opt-out of surgical intervention. This bias was somewhat alleviated by including *‘Sy: Abdominal pain’* to act as the main interdependent agent between symptoms and as the main indicator for a patient to seek medical aid. Considering endometriosis as a possible diagnosis in women who present with abdominal pain may improve its recognition in primary care settings.

The results from the socio-economic section of the model exploit the hidden, or latent, casual variables that account for the lower rate of diagnosis for patients from a black racial background compared with patients identifying as white and Asian. The socio-economic aspect of the model demonstrates that observed racial disparities in health reflect the differences in socio-economic circumstances among racial groups. This information can be used to inform policy-makers of the importance of access to healthcare, regardless of income levels, for the treatment and wellbeing of those who have the condition. The model outputs low levels of quality of life for individuals who have endometriosis but who have not received a formal diagnosis.

### Future Work

The model created is primarily a diagnostic model calculating the probability of a patient having, and subsequently being diagnosed, with endometriosis. It also explores the reality of how endometriosis affects the patient’s quality of life. Whilst treatment probabilities are included, the model is considered a diagnostic rather than an intervention model. To make this a dynamic model it would benefit by being extended to a two-stage model by creating a link between pre-procedure symptoms and post-treatment outcomes. Further research will need to be undertaken to associate range of symptoms with the stage of endometriosis, stage 1-2 (minimal to mild) or stage 3-4 (moderate to severe), as this classification alongside consultation with the patient to ascertain their fertility options and requests will be needed to complete the decision-making process.

The complications and co-morbidities associated with endometriosis can be expanded upon as the model advances. Of interest is the prevalence of mental health issues arising from levels of pain experienced by the patient and the disregard exhibited by clinicians when treating this symptom and subsequent delay of associating it with endometriosis. To be given an explanation for their problems is almost as important as getting a cure for certain individuals (Grundström, et al., 2016).

The model would benefit from a user-centric interface if rolled out to clinicians, to aid decision-making, and policymakers, to illustrate the disadvantages experienced by lower-income workers in their access to medical care.

## Data Availability

The model was constructed using AgenaRisk software (www.agenarisk.com). The model is available for download at www.eecs.qmul.ac.uk/~norman/Models/ (filename endo.cmpx2) and can be run in the free trial version of AgenaRisk.
The model parameters are derived from the data reported in multiple medical observational studies and trials.

http://www.eecs.qmul.ac.uk/~norman/Models/

## Appendix A Model Images

## Appendix B Diagnosis and Treatment results

## Appendix C Race and Ethnicity factors

## Appendix D Satisfaction with care

